# Trends in types of revision for prosthetic joint infection and risk of a second revision from 1995 to 2022: A descriptive cohort study from the Danish Hip Arthroplasty Registry

**DOI:** 10.1101/2024.01.04.24300764

**Authors:** Daniel Lundsgaard, Armita Armina Abedi, Søren Overgaard, Jens Holm Laigaard

**Affiliations:** Department of Orthopedic surgery, Bispebjerg and Frederiksberg Hospital; Department of Clinical Medicine, Faculty of Health and Medical Sciences, University of Copenhagen, Copenhagen, Denmark

**Keywords:** Joint Revision, Debridement, Hip replacement, Hip prosthesis, Prosthesis-Related Infections

## Abstract

This protocol outlines a register-based cohort study utilizing Danish Hip Arthroplasty Register data from 1995 to 2022. Focusing on adult patients undergoing primary total hip arthroplasty (THA), the study categorizes revisions, particularly periprosthetic joint infections (PJI), into Debridement, antibiotics, and implant retention (DAIR), one-stage, and two-stage approaches. Annual revision numbers and the risk of a second revision after each primary type will be analyzed. The study’s strengths lie in its comprehensive examination of various PJI revision methods and an extended recording period, leveraging the DHR’s high completeness rates. While acknowledging potential uncertainties, the study offers valuable insights into THA revision trends, aiding in clinical practice optimization and improving patient outcomes.

**Study registration:** Privacy identifier p-2023-14990

Protocol uploaded to OSF: Not yet uploaded

**Budget:** The costs of the study are limited to the salary of the investigators, which is paid by the orthopedic department.

## Rationale and background

Total hip arthroplasty (THA) is one of the most used surgical procedures and has been increasing during several years, and according to projections it will increase further in the coming years.^1^ Over $1.3 billion (CAD) was spent on hospitalizations for hip and knee replacement surgeries in Canada from 2020 to 2021, and in Australia the predictions are that in 2030, the number will exceed 5.32 billion dollars, the economic impact is thereby a big factor in talking THA operations.^1^ Although it is commonly a successful procedure, there is a risk of complications, which include periprosthetic joint infection (PJI). According to different studies PJI has an upgoing trend, and has risen a significant amount in the latest years. ^2 3 4^ This shows an unfortunate increase of revisions caused by infections. The upgoing trend can either reflect a true increase (frailer patients or more use of uncemented implants) or an apparent increase (improved diagnostics, more reporting or changed in revision strategy).^3^ PJI’s can be devastating for the patients as it causes emotional destress, can result in functional disability, and in a some cases, lead to a fatal outcome.^5^ In this study PJI is considered present when the following criteria exists: A PJI when an indication of deep infection is reported to DHR by the surgeon upon revision surgery.^2^

Different approaches to revision surgery can be used to treat PJI. These approaches can be categorized as Debridement, antibiotics, and implant retention (DAIR), one-stage revision and two-stage revision. Which approach is chosen depends on several factors, such as duration of symptoms, the infectious agent, and the patient’s comorbidities. Ideally, this choice is made by a combination of an experienced orthopedic surgeon, an infectious disease specialist and a well-informed patient.

For acute and hematogenous infections, the preferred approach is often DAIR. DAIR aims at addressing PJIs without replacing non-modular components, thus preserving the integrity of the implant and the function of the joint. It is normaly used when the infection is caught early and has not caused significant damage to the surrounding tissue around the implant. DAIR is defined as an exchange of the modular parts after a THA. The exchange can be of acetabular liner, femoral head, the neck segment and soft tissue replacement without replacement of prosthetic components.

In chronic PJI or later discovered PJIs where biofilm, multi-drug resistant bacteria or fungi complicates the infection,^6^ patients are typically offered 1-stage- or a 2-stage revision. In a 1-stage revision all device components are removed, and new revision components are inserted after debridement as part of the same surgical procedure. In a 2-stage revision, all the components are removed in the first stage, and a temporary antibiotic-laden spacer device is placed. The second stage, definite reimplantation, is typically performed after a long course of antibiotics. Spacers were historically composed entirely of antibiotic cement (e.g, polymethyl methacrylate (PMMA)), but there has been an increase in spacers compromised of a combination of PMMA, metal and polyethylene, which provide a smoother surface, similar to the permanent implants.^6^

While 2-stage exchange arthroplasty is generally preferred for treatment of chronic PJI, some studies have challenged this practice by showing non-inferior results with 1-stage exchange surgery.^7 8^ According to a 2021 meta-analysis, 70% of all DAIR procedures in hip PJIs were successful.^4^ In addition, the study found that the success rate increased over time. There was a 2.6% increase in success rate for each 10% increase in component exchange. However, when restricted to studies after 2004, the increase in component exchange didn’t make a difference.^4^ Two systematic reviews have shown that the success rate of DAIRs has improved over the last 20 years, and one explanation could be newer and more effective antibiotic treatment, surgical techniques and the timing of DAIR.^9 10^

According to various articles, there is significant uncertainty in the diagnosis and treatment of PJI. Many factors need to be considered, making it incredibly challenging to achieve the most accurate outcomes. Through a period of time there has been a clarification process underway about the international guidelines to establish the most accurate guidelines going forward. This has resulted in a healthy debate according PJI, which has affected the view on PJI diagnose and treatment. The question then becomes whether these new thoughts on guidelines has been implemented in practice.^11,12^

We therefore aim is to describe trends in the types of revision in PJI following primary THA, categorized into DAIR, one-stage exchange and two-stage implant exchange. Further, we aim to describe the risk of a second revision, following each of these revision types.

## Methods

This protocol was written in accordance with the Strengthening the Reporting of Observational Studies in Epidemiology (STROBE) guideline for reporting of cohort studies^13,14^, aside from the results section, as this is a protocol.

### Study design

This is a protocol for a descriptive, register-based, cohort study.

### Setting

All residents in Denmark have equal access to free-of-charge healthcare, including THA, which is financed by general taxes. The majority of patients undergo THA and THA revision surgery at public hospitals, though the proportion of surgeries conducted at private hospitals is increasing.^2^

We will include data from all public and private orthopedic departments from 1995 to 2022. The data collection will start on the day of the primary THA operation and continue until death or the date of first revision, whichever occurs first. We will consider the entire available follow-up period for all patients, i.e., up to 28 years after primary THA surgery. All patients are followed up for at least 90 days to ensure correct categorization of the time between primary and revision surgery (see the quantitative variables section).

### Participants

Inclusion criteria: All adult (≥18 years old) patients who underwent primary THA for any reason during the study period are eligible. Patients/hips enter the study on the day of their primary THA surgery. A patient can enter the study twice – once for each hip.

Exclusion criteria: Patients who underwent hemiarthroplasty or simultaneous bilateral arthroplasty are excluded.

### Data sources

All data used in this study was found in the Danish Hip Arthroplasty Register (DHR). The DHR collects data from all THAs performed in Denmark, including both primary THAs and revisions. It is mandatory for all surgeons performing THAs to report to the register, which results in high completeness. The data in DHR is continuously validated with data from the Danish Patient Register. Further, studies have shown that the majority of PJI are correctly reported.^16^,^17^. In the case of a revision, the surgeon reports the indication immediately after surgery. The revisions performed from 1995-2020 are located in the DHR, which is enriched with data from the Danish patient Registry.^18^,^19^

### Quantitative variables

We will present the total and yearly number of patients who underwent revision surgery for any reason, as well as the number of revisions from each revision cause (percentage of all revisions).

As the primary focus of this study, we will present the yearly number of revision surgeries for PJI overall, and divided into the three surgical approaches: DAIR, one-stage exchange revision and two-stage exchange revision (percentage of all revisions due to PJI). The numbers of revision surgeries for PJIs will also be analyzed in three subgroups, depending on the time between primary and revision surgery: ^20^

a. Within 4 weeks after primary surgery
b. Between 4 and 12 weeks after primary surgery
c. More than 12 weeks after primary surgery

Finally, we will report the risk of a second revision after each type of surgical approach.

### Statistical methods

To ease data management and reduce the risk of errors, the patients will be analyzed by THA side (right/left). All data will be analyzed in the newest version of R statistical software^21^ with packages tidyverse and epitools^22^,^23^.

The analysis of risk of second revision after the different types of primary revisions, will be performed using the Survival package in R with cox proportional hazards model, adjusted for calendar time, age and sex.

## Results

We will report the study results according to the STROBE statement, including a diagram of patients’ (hips’) flow through the study. The risk of a second revision for each type of primary revisions, will be presented as a Kaplan Meier plot.

### Strengths and limitations

Some of the strengths of this article include the examination of data for various PJI revision methods, which will provide a more comprehensive perspective on our data compared to other articles that have focused solely on either DAIR or only 1-stage, 2-stage revision. Additionally, by looking at data dating back to 1995, a long recording period is achieved, which we believe is necessary to present trends in surgeons’ practice.

DHR has a completeness rate of 96.9% for primary total hip arthroplasty (THA) and 92.3% for revisions. It is observed that 7 out of 16 private clinics do not have satisfactory completeness, which introduces some uncertainties in DHR’s data. However, DHR meets the regulatory requirement of 90% completeness. PJIs are surgeon-reported and not necessarily verified by paraclinical findings. Thus, it may cause misdiagnosed patients to be recorded as PJI revisions. Still, as we are interested in surgeons’ practice, we consider the actual diagnosis of lesser importance.

The prognosis, including the results of the cox model, should be viewed only as descriptive figures. This is an observational study, and the aim of the study is descriptive.

### Generalizability

Our study relies on data from Denmark only. The figures, trends, and prognosis may therefore be different in other settings, and especially in non-developed countries. Still, we use very broad inclusion criteria and therefore, the study population closely resembles the clinical population. We believe the figures presented in our study, may be similar to those in other developed countries.

## Data Availability

All data produced in the present study can be obtained from Danish Hip Register.

